# Comparison of COVID-19 outcomes among shielded and non-shielded populations: A general population cohort study of 1.3 million

**DOI:** 10.1101/2020.09.17.20196436

**Authors:** Bhautesh D Jani, Frederick K Ho, David J Lowe, Jamie P Traynor, Sean MacBride-Stewart, Patrick B Mark, Frances S Mair, Jill P Pell

## Abstract

Many western countries used shielding (extended self-isolation) of people presumed to be at high-risk from COVID-19 to protect them and reduce healthcare demand. To investigate the effectiveness of this strategy, we linked family practitioner, prescribing, laboratory, hospital and death records and compared COVID-19 outcomes among shielded and non-shielded individuals in the West of Scotland. Of the 1.3 million population, 27,747 (2.03%) were advised to shield, and 353,085 (26.85%) were classified a priori as moderate risk. COVID-19 testing was more common in the shielded (7.01%) and moderate risk (2.03%) groups, than low risk (0.73%). Referent to low-risk, the shielded group had higher confirmed infections (RR 8.45, 95% 7.44-9.59), case-fatality (RR 5.62, 95% CI 4.47-7.07) and population mortality (RR 57.56, 95% 44.06-75.19). The moderate-risk had intermediate confirmed infections (RR 4.11, 95% CI 3.82-4.42) and population mortality (RR 25.41, 95% CI 20.36-31.71) but, due to their higher prevalence, made the largest contribution to deaths (PAF 75.30%). Age ≥70 years accounted for 49.55% of deaths. In conclusion, shielding has not been effective at preventing deaths in individuals at high risk. Also, to be effective as a population strategy, shielding criteria would need to be widely expanded to include other criteria, such as the elderly.

## Introduction

Early in the COVID-19 pandemic, a major concern was that the demand on health services would exceed capacity in terms of hospitalisations, intensive care unit (ICU) admissions, and ventilation ^1^. It was assumed that sub-groups of the population would have worse prognosis and, therefore, contribute disproportionately to adverse outcomes and healthcare demands.

Asian countries generally relied on population-wide strategies ^2^. Early, widespread ‘test, trace, isolate’ strategies were made possible by higher testing capacity and greater willingness to monitor and enforce compliance. In contrast, Europe and the USA adopted a two-pronged approach; ^2^ general population interventions, such as physical distancing and hand hygiene, designed to reduce transmission in the population as a whole, supplemented by shielding of those assumed to be at higher risk. Notably, Sweden, an outlier in not applying lock-down, nonetheless mandated shielding^3^.

In the UK, a Vulnerable Patient List (Supplementary Table 1)^6^ was produced comprising two categories labelled high risk, highest risk or clinically extremely vulnerable and moderate risk, at risk or clinically vulnerable by various UK organisations. In this manuscript, they are referred to as shielded and moderate risk respectively, with the remaining population labelled low-risk. In the UK, the shielded group received individual letters strongly recommending they self-isolate over a protracted period - not leaving their homes and avoiding non-essential contact with household members - and were provided with support at home such as delivery of food packages. The moderate risk category was simply advised to be vigilant in adhering to general advice.

The category definitions were based largely on expert opinion informed by our understanding of previous viruses and the need for better definitions has been highlighted ^7^. Studies are emerging of the risk factors associated with COVID-19 outcomes. Among two million UK community-based app users self-reported heart disease, kidney disease, lung disease, diabetes and obesity were associated with self-reported hospital admission and respiratory support for COVID-19 ^8^. Similarly, linkage of family practitioner records of 17 million people in England reported a wide range of long-term conditions associated with in-hospital death from COVID-19 including: respiratory, heart, liver and kidney disease, diabetes, cancers, stroke and organ transplantation ^9^. Unfortunately, the investigators did not have access to deaths in the community. COVID-19 risk scores are being developed in an attempt to improve identification of high risk individuals who could be advised to shield ^10^ but attempts to investigate the potential contribution of a shielding strategy to population-level outcomes and healthcare demands have so far been limited to mathematical modelling^11–19^.

The aims of this study were to compare those classified, a priori, as high risk (and therefore advised to shield) and those classified as moderate and low-risk, in terms of their individual risk of COVID-19 infection and outcomes and the extent to which they accounted for COVID-19 related outcomes at a population level.

## Results

Of the 1,315,071 people registered with family practitioners in NHS Greater Glasgow and Clyde in the West of Scotland, 26,747 (2.03%) were on the shielding list and 353,085 (26.85%) were classified, a priori, as moderate-risk. Of the 26,747 shielded group, 18,147 (55.78%) had severe respiratory disease, 5,349 (16.44%) were on immunosuppressive therapies, 2,491 (7.66%) had specific cancers, 1,245 (3.83%) had received organ transplants, 475 (1.78%) were on renal dialysis, and less than five were pregnant and had severe heart disease. Of the 353,085 classified as moderate-risk, 160,215 (45.38%) had hypertension, 151,865 (43.01%) had chronic lung disease, 139,568 (39.53%) were ≥70 years of age, 64,358 (18.23%) had diabetes, 48,571 (13.81%) had heart disease, and 1,195 (0.34%) had a weakened immune system.

### Shielded and moderate-risk categories

Overall, 15,865 (1.21%) people were tested for COVID-19. The likelihood of being tested increased with age, was higher in women and the moderate-risk category and highest in the shielded group (Table 1). Overall, 3,348 (0.25%) people had confirmed COVID-19 infection. The likelihood of laboratory-confirmed COVID-19 infection followed similar patterns as testing. It increased with age, was higher in women, was highest in the shielded group and lowest in the low-risk category (Table 2). After adjustment for sex and deprivation quintile, the risk of laboratory-confirmed infection remained higher in the moderate-risk category and highest in the shielded group (Table 3).

**Table 1.** COVID-19 testing status by sociodemographic characteristics, risk category and risk criteria

|  | COVID-19 testing status |  | P value |
| --- | --- | --- | --- |
|  | Not tested<br>N=1,299,206<br>n (%) | Tested<br>N=15,865<br>n (%) |  |
| Age group (years) |  |  | <0.0001 |
| 0-24 | 355,238 (99.49) | 1,822 (0.51) |  |
| 25-44 | 410,408 (99.22) | 3,247 (0.78) |  |
| 45-64 | 340,268 (98.65) | 4,660 (1.35) |  |
| ≥65 | 193,292 (96.92) | 6,136 (3.08) |  |
| Sex |  |  | <0.0001 |
| Male | 654,041 (99.01) | 6,569 (0.99) |  |
| Female | 645,165 (98.58) | 9,296 (1.42) |  |
| Deprivation quintile |  |  | <0.0001 |
| 1 (most deprived) | 461,672 (98.67) | 6,211 (1.33) |  |
| 2 | 230,402 (98.75) | 2,921 (1.25) |  |
| 3 | 194,702 (98.90) | 2,175 (1.10) |  |
| 4 | 173,456 (98.93) | 1,883 (1.07) |  |
| 5 (most affluent) | 238,974 (98.89) | 2,675 (1.11) |  |
| Risk category |  |  | <0.0001 |
| Low | 928,420 (99.27) | 6,819 (0.73) |  |
| Moderate | 345,913 (97.97) | 7,172 (2.03) |  |
| Shielded | 24,873 (92.99) | 1,874 (7.01) |  |
| Moderate risk criteria |  |  |  |
| Chronic respiratory disease | 149,325 (98.33) | 2,540 (1.67) | <0.0001 |
| Heart disease | 46,728 (96.21) | 1,843 (3.79) | <0.0001 |
| Hypertension | 156,286 (97.55) | 3,929 (2.45) | <0.0001 |
| Diabetes | 62,482 (97.09) | 1,876 (2.91) | <0.0001 |
| Weakened immune system | 1,140 (95.40) | 55 (4.60) | <0.0001 |
| ≥70 years of age | 134,305 (96.23) | 5,263 (3.77) | <0.0001 |
| Shielded criteria |  |  |  |
| Severe respiratory disease | 17,146 (94.48) | 1,001 (5.52) | <0.0001 |
| Specific cancers | 2,075 (83.30) | 416 (16.70) | <0.0001 |
| Pregnant with severe heart disease | <5 | 0 | - |
| Immunosuppressive therapy | 5,028 (94.00) | 321 (6.00) | <0.0001 |
| Solid organ transplant | 1,149 (92.29) | 96 (7.71) | <0.0001 |
| Rare diseases and inborn errors of metabolism | 1,623 (91.95) | 142 (8.05) | <0.0001 |
| Renal dialysis | 305 (64.21) | 170 (35.79) | <0.0001 |

N number

**Table 2.** Crude, population-level COVID-19 outcomes by sociodemographic characteristics, risk category and risk criteria

|  | Confirmed COVID-19 infection |  |  | COVID-19 hospitalisation |  |  | COVID-19 ICU admission |  |  | COVID-19 mortality |  |  |
| --- | --- | --- | --- | --- | --- | --- | --- | --- | --- | --- | --- | --- |
|  | Negative test/<br>Not tested | Positive test | P-value | Not admitted | Admitted | P-value | Not admitted | Admitted | P-value | Alive | Dead | P-value |
|  | N=1,311,723<br>n (%) | N=3,348<br>n (%) |  | N=1,313,410<br>n (%) | N=1,661<br>n (%) |  | N=1,314,949<br>n (%) | N=122<br>n (%) |  | N=1,314,044<br>n (%) | N=1,027<br>n (%) |  |
| Age group (years) |  |  | <0.0001 |  |  | <0.0001 |  |  | 0.0005 |  |  | <0.0001 |
| 0-24 | 356,944 (99.97) | 116 (0.03) |  | 357,041 (99.99) | 19 (0.01) |  | 357,060 (100.00) | 0 |  | 357,060 (100.00) | 0 (0.00) |  |
| 25-44 | 413,123 (99.87) | 532 (0.13) |  | 413,532 (99.97) | 123 (0.03) |  | 413,643 (100.00) | 12 (0.00) |  | 413,647 (100.00) | 8 (0.00) |  |
| 45-64 | 343,856 (99.69) | 1,072 (0.31) |  | 344,409 (99.85) | 519 (0.15) |  | 344,846 (99.98) | 82 (0.02) |  | 344,838 (99.97) | 90 (0.03) |  |
| ≥65 | 197,800 (99.18) | 1,628 (0.82) |  | 198,428 (99.50) | 1,000 (0.50) |  | 199,400 (99.99) | 28 (0.01) |  | 198,499 (99.53) | 929 (0.47) |  |
| Sex |  |  |  |  |  |  |  |  |  |  |  |  |
| Male | 659,203 (99.79) | 1,407 (0.21) | <0.0001 | 659,781 (99.87) | 829 (0.13) | 0.81 | 660,525 (99.99) | 85 (0.01) | <0.0001 | 660,092 (99.92) | 518 (0.08) | 0.92 |
| Female | 652,250 (99.70) | 1,941 (0.30) |  | 659,369 (99.87) | 832 (0.13) |  | 660,908 (99.99) | 37 (0.01) |  | 653,952 (99.92) | 509 (0.08) |  |
| Deprivation quintile |  |  | 0.0002 |  |  | <0.0001 |  |  | 0.18 |  |  | <0.0001 |
| 1 (most deprived) | 466,582 (99.72) | 1,301 (0.28) |  | 467,146 (99.84) | 737 (0.16) |  | 467,832 (99.99) | 51 (0.01) |  | 467,442 (99.91) | 441 (0.09) |  |
| 2 | 232,710 (99.74) | 613 (0.26) |  | 233,041 (99.88) | 282 (0.12) |  | 233,302 (99.99) | 21 (0.01) |  | 233,181 (99.94) | 142 (0.06) |  |
| 3 | 196,416 (99.77) | 461 (0.23) |  | 196,651 (99.89) | 226 (0.11) |  | 196,854 (99.99) | 23 (0.01) |  | 196,724 (99.92) | 153 (0.08) |  |
| 4 | 174,939 (99.77) | 400 (0.23) |  | 175,143 (99.89) | 196 (0.11) |  | 175,329 (99.99) | 10 (0.01) |  | 175,209 (99.93) | 130 (0.07) |  |
| 5 (most affluent) | 241,076 (99.76) | 573 (0.24) |  | 241,429 (99.91) | 220 (0.09) |  | 241,632 (99.99) | 17 (0.01) |  | 241,488 (99.93) | 161 (0.07) |  |
| Risk category |  |  | <0.0001 |  |  | <0.0001 |  |  | <0.0001 |  |  | <0.0001 |
| Low | 934,049 (99.87) | 1,190 (0.13) |  | 93,4839 (99.96) | 400 (0.04) |  | 935,174 (99.99) | 65 (0.01) |  | 935,155 (99.99) | 84 (0.01) |  |
| Moderate | 351,226 (99.47) | 1,859 (0.53) |  | 35,2054 (99.71) | 1,031 (0.29) |  | 353,033 (99.99) | 52 (0.01) |  | 352,282 (99.77) | 803 (0.23) |  |
| Shielded | 26,448 (98.88) | 299 (1.12) |  | 26,517 (99.14) | 230 (0.86) |  | 26,742 (99.98) | 5 (0.02) |  | 26,607 (99.48) | 140 (0.52) |  |
| Moderate risk criteria |  |  |  |  |  |  |  |  |  |  |  |  |
| Chronic respiratory disease | 151,414 (99.70) | 451 (0.30) | 0.0005 | 151,618 (99.84) | 247 (0.16) | <0.0001 | 151,853 (99.99) | 12 (0.01) | 0.65 | 151,812 (99.97) | 53 (0.03) | <0.0001 |
| Heart disease | 48,176 (99.19) | 395 (0.81) | <0.0001 | 48,325 (99.49) | 246 (0.51) | <0.0001 | 48,564 (99.99) | 7 (0.01) | 0.34 | 48,456 (99.76) | 115 (0.24) | <0.0001 |
| Hypertension | 159,267 (99.41) | 948 (0.59) | <0.0001 | 159,670 (99.66) | 545 (0.34) | <0.0001 | 160,189 (99.98) | 26 (0.02) | 0.003 | 159,991 (99.86) | 224 (0.14) | <0.0001 |
| Diabetes | 63,903 (99.29) | 455 (0.71) | <0.0001 | 64,063 (99.54) | 295 (0.46) | <0.0001 | 64,335 (99.96) | 23 (0.04) | <0.0001 | 64,263 (99.85) | 95 (0.15) | <0.0001 |
| Weakened immune system | 1,183 (99.00) | 12 (1.00) | <0.0001 | 1,185 (99.16) | 10 (0.84) | <0.0001 | 1,195 (100.00) | 0 (0.00) | - | 1,189 (99.50) | 6 (0.50) | <0.0001 |
| ≥70 years of age | 138,115 (98.96) | 1,453 (1.04) | <0.0001 | 138,701 (99.38) | 867 (0.62) | <0.0001 | 139,560 (99.99) | 8 (0.01) | 0.19 | 138,690 (99.37) | 878 (0.63) | <0.0001 |
| Shielded group |  |  |  |  |  |  |  |  |  |  |  |  |
| Severe respiratory disease | 17,981 (99.09) | 166 (0.91) | <0.0001 | 18,012 (99.26) | 135 (0.74) | <0.0001 | 18,146 (99.99) | <5 | - | 18,059 (99.52) | 88 (0.48) | <0.0001 |
| Specific cancers | 2,452 (98.43) | 39 (1.57) | <0.0001 | 2,462 (98.84) | 29 (1.16) | <0.0001 | 2,491 (100.00) | 0 | - | 2,475 (99.36) | 16 (0.64) | <0.0001 |
| Pregnant, severe heart disease | <5 | 0 | - | <5 | 0 | - | <5 | 0 | - | <5 | 0(0.00) | - |
| Immunosuppressive therapy | 5,285 (98.80) | 64 (1.20) | <0.0001 | 5,299 (99.07) | 50 (0.93) | <0.0001 | 5,346 (99.94) | <5 | - | 5,324 (99.53) | 25 (0.47) | <0.0001 |
| Solid organ transplant | 1,228 (98.63) | 17 (1.37) | <0.0001 | 1,230 (98.80) | 15 (1.20) | <0.0001 | 1,244 (99.92) | <5 | - | 1,238 (99.44) | 7 (0.56) | <0.0001 |
| Rare diseases and IEM | 1,729 (97.96) | 36 (2.04) | <0.0001 | 1,741 (98.64) | 24 (1.36) | <0.0001 | 1,764 (99.94) | <5 | - | 1,744 (98.81) | 21 (1.19) | <0.0001 |
| Renal dialysis | 445 (93.68) | 30 (6.32) | <0.0001 | 457 (96.21) | 18 (3.79) | <0.0001 | 475 (100.00) | 0 (0.00) | - | 468 (98.53) | 7 (1.47) | <0.0001 |

N number; IEM inborn errors of metabolism

**Table 3.** Associations* between risk categories and risk criteria and population-level COVID-19 outcomes

|  | Confirmed COVID-19 infection |  | COVID-19 hospitalisation |  | COVID-19 ICU admission |  | COVID-19 mortality |  |
| --- | --- | --- | --- | --- | --- | --- | --- | --- |
|  | RR (95% CI) | P-value | RR (95% CI) | P-value | RR (95% CI) | P-value | RR (95% CI) | P-value |
| **Low** | 1 (Reference) |  | 1 (Reference) |  | 1 (Reference) |  | 1 (Reference) |  |
| **Moderate** |  |  |  |  |  |  |  |  |
| Overall | 4.11 (3.83-4.42) | <0.0001 | 6.83 (6.09-7.67) | <0.0001 | 2.15 (1.49-3.10) | <0.0001 | 25.41 (20.36-31.71) | <0.0001 |
| Chronic respiratory disease | 2.30 (2.07-2.56) | <0.0001 | 3.76 (3.21-4.41) | <0.0001 | 1.15 (0.62-2.14) | 0.65 | 3.88 (2.77-5.44) | <0.0001 |
| Heart disease | 6.53 (5.83-7.31) | <0.0001 | 11.63 (9.93-13.63) | <0.0001 | 1.92 (0.88-4.20) | 0.1 | 26.00 (19.71-34.31) | <0.0001 |
| Hypertension | 4.60 (4.22-5.01) | <0.0001 | 8.01 (7.04-9.11) | <0.0001 | 2.44 (1.55-3.85) | 0.0001 | 15.68 (12.24-20.09) | <0.0001 |
| Diabetes | 5.59 (5.02-6.23) | <0.0001 | 10.49 (9.03-12.19) | <0.0001 | 4.89 (3.03-7.89) | <0.0001 | 16.23 (12.16-21.66) | <0.0001 |
| Weakened immune system | 7.79 (4.43-13.72) | <0.0001 | 19.62 (10.50-36.69) | <0.0001 | - | - | 56.19 (24.85-127.02) | <0.0001 |
| ≥70 years of age | 8.08 (7.48-8.72) | <0.0001 | 14.96 (13.29-16.85) | <0.0001 | 0.90 (0.43-1.86) | 0.77 | 72.98 (58.81-90.55) | <0.0001 |
| **Shielded** |  |  |  |  |  |  |  |  |
| Overall | 8.45 (7.44-9.59) | <0.0001 | 19.35 (16.45-22.77) | <0.0001 | 2.78 (1.12-6.91) | 0.03 | 57.56 (44.06-75.19) | <0.0001 |
| Severe respiratory disease | 6.79 (5.78-7.99) | <0.0001 | 16.40 (13.48-19.95) | <0.0001 | 0.82 (0.11-5.89) | 0.84 | 52.85 (39.31-71.05) | <0.0001 |
| Specific cancers | 12.13 (8.83-16.65) | <0.0001 | 27.21 (18.69-39.63) | <0.0001 | - | - | 71.72 (42.36-121.42) | <0.0001 |
| Immunosuppressive therapy | 9.25 (7.21-11.88) | <0.0001 | 21.85 (16.30-29.29) | <0.0001 | 8.40 (2.64-26.72) | 0.0003 | 52.23 (33.64-81.08) | <0.0001 |
| Solid organ transplant | 10.90 (6.77-17.55) | <0.0001 | 27.77 (16.61-46.44) | <0.0001 | 10.84 (1.50-78.16) | 0.02 | 62.03 (29.03-132.56) | <0.0001 |
| Rare diseases and IEM | 15.91 (11.44-22.12) | <0.0001 | 31.45 (20.86-47.42) | <0.0001 | 8.18 (1.13-58.93) | 0.04 | 132.29 (82.60-211.86) | <0.0001 |
| Renal dialysis | 50.29 (35.06-72.12) | <0.0001 | 84.41 (52.70-135.20) | <0.0001 | - | - | 158.41 (74.11-338.62) | <0.0001 |

\*adjusted for sex and deprivation quintile
RR relative risk; CI confidence interval; IEM inborn errors of metabolism

Overall, 1,661 people were hospitalised for COVID-19. Within the general population, hospitalisations increased with age but were comparable between men and women (Table 2). Hospitalisations were more common in the moderate-risk category and most common in the shielded group (Table 2), remaining so after adjustment for sex and deprivation (Table 3). Overall, 122 people were admitted to ICU wards for COVID-19. ICU admissions were significantly more common among people aged 45-64 years of age than among older people (Table 2). Compared with the low-risk category, the shielded group were 18 times more likely to be hospitalised but only 4 times more likely to be admitted to ICU (Table 3). Overall, 1,027 (0.08%) people died from COVID-19. Within the general population, mortality increased with age but was similar in men and women (Table 2). Population mortality was higher in the moderate-risk category and highest in the shielded group (Table 2) and remained so after adjustment for sex and deprivation (Table 3).

Among the sub-group with laboratory-confirmed (test-positive) COVID-19 infection, 1,661 (49.6%) were hospitalised. Hospitalisations increased with age but were comparable between men and women (Table 4). The moderate-risk category was more likely to be hospitalised and the shielded group most likely (Table 4), remaining so after adjustment for age and deprivation (Table 5). Among those with laboratory-confirmed infection, ICU admissions were more common in men and more common in people aged 45-64 years than those older (Table 4). Low-risk cases were more likely to be admitted to ICU than the moderate-risk and shielded groups (Tables 4 & 5). Among the sub-group with clinically-confirmed (test-positive or COVID-19 related death) COVID-19 infection, 1,027 (26.70%) died (Table 4). Case-fatality increased by age and was higher in men than women. It was lowest in the low-risk category but not significantly different between the moderate-risk and shielded groups (RR_shielded/moderate_ [95% CI] 1.12 [0.96-1.31], p=0.14) (Table 5).

**Table 4.** Crude COVID-19 outcomes among confirmed cases by sociodemographic characteristics, risk category and risk criteria

|  | COVID-19 hospitalisation<br>N=3,348<sup>†</sup> |  |  | COVID-19 ICU admission<br>N=3,348<sup>†</sup> |  |  | COVID-19 case-fatality<br>N=3,846<sup>‡</sup> |  |  |
| --- | --- | --- | --- | --- | --- | --- | --- | --- | --- |
|  | Not admitted<br>N=1,687<br>n (%) | Admitted<br>N=1,661<br>n (%) | P-value | Not admitted<br>N=3,226<br>n (%) | Admitted<br>N=122<br>n (%) | P-value | Alive<br>N=2,819<br>n (%) | Dead<br>N=1,027<br>n (%) | P-value |
| Age group (years) |  |  | <0.0001 |  |  | <0.0001 |  |  | <0.0001 |
| 0-24 | 97 (83.62) | 19 (16.38) |  | 116 (100.00) | 0 |  | 116 (100.00) | 0 (0.00) |  |
| 25-44 | 410 (76.92) | 123 (23.08) |  | 520 (97.74) | 12 (2.26) |  | 526 (98.50) | 8 (1.50) |  |
| 45-64 | 553 (51.59) | 519 (48.41) |  | 990 (92.35) | 82 (7.65) |  | 1,003 (91.77) | 90 (8.23) |  |
| ≥65 | 630 (38.65) | 1,000 (61.35) |  | 1,600 (98.28) | 28 (1.72) |  | 1,174 (55.83) | 929 (44.17) |  |
| Sex |  |  |  |  |  |  |  |  | <0.0001 |
| Male | 579 (41.12) | 829 (58.88) | <0.0001 | 1,322 (93.96) | 85 (6.04) | <0.0001 | 1,105 (68.08) | 518 (31.92) |  |
| Female | 1,108 (57.11) | 832 (42.89) |  | 1,904 (98.09) | 37 (1.91) |  | 1,717 (77.10) | 509 (22.90) |  |
| Deprivation Quintile |  |  | <0.0001 |  |  | 0.29 |  |  | 0.0004 |
| 1 (most deprived) | 566 (43.44) | 737 (56.56) |  | 1,250 (96.08) | 51 (3.92) |  | 1,046 (70.34) | 441 (29.66) |  |
| 2 | 332 (54.07) | 282 (45.93) |  | 592 (96.57) | 21 (3.43) |  | 531 (78.90) | 142 (21.10) |  |
| 3 | 235 (50.98) | 226 (49.02) |  | 438 (95.01) | 23 (4.99) |  | 394 (72.03) | 153 (27.97) |  |
| 4 | 204 (51.00) | 196 (49.00) |  | 390 (97.50) | 10 (2.50) |  | 341 (72.40) | 130 (27.60) |  |
| 5 (most affluent) | 353 (61.61) | 220 (38.39) |  | 556 (97.03) | 17 (2.97) |  | 507 (75.90) | 161 (24.10) |  |
| Risk category |  |  | <0.0001 |  |  | 0.0001 |  |  | <0.0001 |
| Low | 791 (66.41) | 400 (33.59) |  | 1125 (94.54) | 65 (5.46) |  | 1130 (93.08) | 84 (6.92) |  |
| Moderate | 828 (44.54) | 1031 (55.46) |  | 1807 (97.20) | 52 (2.80) |  | 1485 (64.90) | 803 (35.10) |  |
| Shielded | 71 (23.59) | 230 (76.41) |  | 294 (98.33) | 5 (1.67) |  | 204 (59.30) | 140 (40.70) |  |
| Moderate risk criteria |  |  |  |  |  |  |  |  |  |
| Chronic respiratory disease | 205 (45.35) | 247 (54.65) | 0.02 | 439 (97.34) | 12 (2.66) | 0.29 | 420 (88.79) | 53 (11.21) | <0.0001 |
| Heart disease | 149 (37.72) | 246 (62.28) | <0.0001 | 388 (98.23) | 7 (1.77) | 0.049 | 330 (74.16) | 115 (25.84) | 0.7 |
| Hypertension | 404 (42.57) | 545 (57.43) | <0.0001 | 922 (97.26) | 26 (2.74) | 0.10 | 836 (78.87) | 224 (21.13) | <0.0001 |
| Diabetes | 161 (35.31) | 295 (64.69) | <0.0001 | 432 (94.95) | 23 (5.05) | 0.11 | 398 (80.73) | 95 (19.27) | <0.0001 |
| Weakened immune system | 2 (16.67) | 10 (83.33) | 0.04 | 12 (100.00) | 0 | - | 6 (50.00) | 6 (50.00) | 0.13 |
| ≥70 years of age | 587 (40.37) | 867 (59.63) | <0.0001 | 1,445 (99.45) | 8 (0.55) | <0.0001 | 1,035 (54.10) | 878 (45.90) | <0.0001 |
| Shielding criteria |  |  |  |  |  |  |  |  |  |
| Severe respiratory disease | 33 (19.64) | 135 (80.36) | <0.0001 | 165 (99.40) | <5 | - | 111 (55.78) | 88 (44.22) | <0.0001 |
| Specific cancers | 10 (25.64) | 29 (74.36) | 0.003 | 39 (100.00) | 0 | - | 27 (62.79) | 16 (37.21) | 0.16 |
| Pregnant, severe heart disease | - | - | - | - | - | - | - | - | - |
| Immunosuppressive therapy | 14 (21.88) | 50 (78.12) | <0.0001 | 61 (95.31) | <5 | - | 42 (62.69) | 25 (37.31) | 0.07 |
| Solid organ transplant | 2 (11.76) | 15 (88.24) | 0.003 | 16 (94.12) | <5 | - | 11 (61.11) | 7 (38.89) | 0.37 |
| Rare diseases and IEM | 12 (33.33) | 24 (66.67) | 0.06 | 35 (97.22) | <5 | - | 23 (52.27) | 21 (47.73) | 0.003 |
| Renal dialysis | 12 (40.00) | 18 (60.00) | 0.33 | 30 (100.00) | 0 | - | 23 (76.67) | 7 (23.33) | 0.83 |

<sup>†</sup>laboratory-confirmed (test-positive) COVID-19 cases<sup>‡</sup>clinically-confirmed (test-positive or COVID-19 on death certificate) COVID-19 cases
N number; IEM inborn errors of metabolism

**Table 5.** Associations* between risk categories and risk criteria and COVID-19 outcomes among confirmed cases

|  | COVID-19 hospitalisation<br>N=3,348<sup>†</sup> |  | COVID-19 ICU admission<br>N=3,348<sup>†</sup> |  | COVID-19 case-fatality<br>N=3,846<sup>‡</sup> |  |
| --- | --- | --- | --- | --- | --- | --- |
|  | RR (95% CI) | P-value | RR (95% CI) | P-value | RR (95% CI) | P-value |
| **Low** | 1 (Reference) |  | 1 (Reference) |  | 1 (Reference) |  |
| **Moderate** |  |  |  |  |  |  |
| Overall | 1.34 (1.23-1.46) | <0.0001 | 0.40 (0.28-0.58) | <0.0001 | 5.01 (4.14-6.06) | <0.0001 |
| Chronic respiratory disease | 1.55 (1.38-1.75) | <0.0001 | 0.46 (0.25-0.86) | 0.02 | 1.61 (1.21-2.16) | 0.001 |
| Heart disease | 1.59 (1.41-1.79) | <0.0001 | 0.24 (0.11-0.53) | 0.0004 | 3.55 (2.79-4.50) | <0.0001 |
| Hypertension | 1.53 (1.39-1.69) | <0.0001 | 0.44 (0.28-0.70) | 0.0004 | 3.04 (2.45-3.76) | <0.0001 |
| Diabetes | 1.69 (1.51-1.90) | <0.0001 | 0.72 (0.45-1.17) | 0.19 | 2.62 (2.05-3.36) | <0.0001 |
| Weakened immune system | 2.41 (1.50-3.86) | 0.0003 | - | - | 7.09 (3.52-14.28) | <0.0001 |
| ≥70 years of age | 1.36 (1.24-1.48) | <0.0001 | 0.07 (0.04-0.15) | <0.0001 | 6.53 (5.42-7.88) | <0.0001 |
| **Shielded** |  |  |  |  |  |  |
| Overall | 1.89 (1.67-2.13) | <0.0001 | 0.23 (0.09-0.56) | 0.001 | 5.62 (4.47-7.07) | <0.0001 |
| Severe respiratory disease | 1.92 (1.66-2.23) | <0.0001 | - | - | 6.16 (4.78-7.93) | <0.0001 |
| Specific cancers | 1.88 (1.41-2.49) | <0.0001 | - | - | 5.00 (3.18-7.87) | <0.0001 |
| Immunosuppressive therapy | 2.15 (1.73-2.69) | <0.0001 | - | - | 5.15 (3.53-7.51) | <0.0001 |
| Solid organ transplant | 2.31 (1.57-3.41) | <0.0001 | - | - | 5.20 (2.71-9.99) | <0.0001 |
| Rare diseases and IEM | 1.49 (1.09-2.03) | 0.01 | - | - | 6.59 (4.39-9.89) | <0.0001 |
| Renal dialysis | 1.61 (1.13-2.30) | 0.009 | - | - | 3.10 (1.61-5.96) | 0.0007 |

\*adjusted for sex and deprivation quintile
<sup>†</sup>laboratory-confirmed (test-positive) COVID-19 cases
<sup>‡</sup>clinically-confirmed (test-positive or COVID-19 on death certificate) COVID-19 cases
N number; RR relative risk; CI confidence interval; IEM inborn errors of metabolism

The shielded group accounted for 7.62% of laboratory-confirmed COVID-19 infections, 12.70% of COVID-19 hospitalisations, 2.69% of ICU admissions and 13.22% of COVID-19 related deaths (Supplementary Table 2). The corresponding figures for the moderate-risk category were 42.06%, 53.28%, 22.96% and 75.30%. To prevent at least 80% of deaths, 28.8% of the population would have had to receive the current level of shielding including those with five criteria classified as moderate-risk at the time of the study (Supplementary Figure 1).

### Individual risk criteria

Due to insufficient numbers, the individual risk criteria models could not be run for pregnant women with severe heart disease or for COVID-19 related ICU admission in the shielded category. All the remaining individual risk criteria were associated with higher likelihood of being tested for COVID-19 (Table 1), laboratory-confirmed infection (Table 2), hospitalisation, population mortality (Table 3) and case-fatality (Table 5) independent of sex and deprivation. Among the moderate-risk category criteria, age ≥70 years and weakened immune system had risks of population mortality (Table 3) and case-fatality (Table 5) at least as high as the overall shielded group. Apart from the 0.13% of people with relevant rare diseases or inborn errors of metabolism and 1.78% on renal dialysis, the strongest associations were observed for those aged ≥70 years who were eight times as likely to have confirmed infection (Table 3); seven times as likely to die following confirmed infection (Table 5); and 74 times as likely to die overall (Table 3) compared with the low-risk category. Being ≥70 years of age accounted for 17.81% of confirmed COVID-19 infections, 22.19% of COVID-19 related hospitalisations, and 49.55% of COVID-19 related deaths (Supplementary Table 2). Among those hospitalised for COVID-19, the likelihood of ICU admission was significantly lower for all individual risk criteria in the moderate-risk category, other than diabetes (Table 5). In particular, hospitalised patients ≥70 years of age were 14 times less likely to be admitted to ICU than low-risk hospitalised patients (Table 5).

## Discussion

The 2.03% of people advised to shield were, nonetheless, eight times more likely to have confirmed infections than the low-risk category, five times more likely to die following confirmed infection and 49 times more likely to die from COVID-19 overall. Whilst selective testing might explain the first outcome, it does not explain higher overall mortality which suggests that the shielding strategy was not as effective as was hoped.

One quarter of the population were classified as moderate-risk and not advised to shield. Nonetheless, they were four times more likely to have confirmed infections than the low-risk category, five times more likely to die following confirmed infection and 25 times more likely to die overall, suggesting that the shielding criteria should be expanded. In particular, older age needs to be considered since the elderly are both at high individual risk and contribute significantly to population burden due to their relatively high numbers.

In spite of people in the shielded and moderate-risk categories having poorer prognosis, they were less likely to be admitted to ICU following hospitalisation for COVID-19, especially patients ≥70 years. This finding reinforces the importance of protection in those with the worst prognosis.

Our finding that 26.85% of people satisfy moderate-risk criteria is consistent with limited existing evidence. A study linking English primary and secondary care records on 3.9 million people reported that 20% of population satisfied similar criteria^20^. Similarly, analysis of the Global Burden of Diseases Study estimated that 22% of the global population are at increased risk of severe COVID-19 disease^21^. A USA study using data from the Behavioral Risk Factor Surveillance System reported that 45.4% of 444,649 adults had one or more of a longer list of morbidities that may be associated with higher risk from COVID-19^22^. Another USA study estimated that 14.2% of participants in the National Health Interview Survey had more than two-fold risk and 1.6% had more than 10-fold risk^23^.

The evidence on COVID-19 related complications among those classified as high risk, and therefore advised to shield, has mainly come from case series and expert opinion. Case series found higher COVID-19 related complications among organ transplant recipients^24,25^, patients receiving chemotherapy, radiotherapy or immunotherapy for cancer^26,27^, and patients with haematological cancers^28^. Systematic review suggested higher COVID-19 complication risk among COPD patients, but the effect of COPD severity was not investigated^29^. Patients with cystic fibrosis and sickle cell disease were classified as high risk based on expert opinion^30,31^. While pregnant women with COVID-19 were found to have higher risk of poor maternal and perinatal outcomes^32,33^, outcomes were not investigated specifically for pregnant women with heart disease. There was no evidence of worse COVID-19 related complications among patients on immunosuppressants ^34^. A large community study in England found strong association between severe asthma (hazard ratio 1.25) and COVID-19 related mortality but did not investigate the risk of COVID-19 infection or hospitalisation^9^.

In common with previous studies, we demonstrated that age was a major individual-level risk factor for death. Additionally, we showed it is important at the population level with 49.55% of deaths attributable to age ≥70 years. The higher mortality in the elderly was mediated in part by higher case-fatality but they also had a higher incidence of infection, possibly due to transmission within care homes. Lower ICU admissions following hospitalisation for COVID-19 may have contributed to their higher case-fatality. Previous studies have reported that men are at higher risk of COVID-19.^7^ Our study demonstrated they are less likely to be tested for COVID-19, have confirmed infection, and be hospitalised. They have comparable overall mortality from COVID-19, due to their lower incidence, but their case-fatality is higher.

This study adds to the existing evidence of the possible effectiveness of a shielding strategy which is currently limited to mathematical modelling of population effects based on assumptions ^11–19^. Ours was a large-scale, unselected general population study. The data cover a period when shielding was in place. Linkage of family practitioner, laboratory, hospital and death data enabled us to examine a range of COVID-19 outcomes and study a range of exposure variables including the overall risk categories and their individual criteria. The datasets were linked using exact, rather than probabilistic, matching. We were able to adjust for potential sociodemographic confounders. The exposure data were collected prior to the outcomes occurring avoiding potential reverse causation and recall or recording bias. Our analysis of potential risk factors was restricted to those used as criteria for shielding and moderate-risk at the time of the study. The shielding and moderate-risk criteria were correct at the time of extracting data but may be revised over time.

Our findings suggest that our attempts to shield those at highest risk have not been as successful as hoped, with those advised to shield experiencing higher rates of infection and death. Since this group was also less likely to be admitted to ICU, protecting them from infection is essential. For shielding to be effective as a population level strategy, the current criteria would need to be expanded since three-quarters of deaths were associated with moderate-risk criteria for which shielding has not hitherto been recommended. In our study, more than one-quarter of the general population would have needed to be effectively shielded to prevent over 80% of deaths. Since this is unlikely to be acceptable at a time when governments are under pressure to avoid further lock-downs, shielding is probably best viewed as an individual-level intervention to be used alongside other population-wide interventions such as physical distancing, face coverings and hand hygiene.

## Methods

We conducted a general population cohort study of all 1.3 million residents of NHS GGC in the West of Scotland. The Community Health Index (CHI), a unique identifier attached to all Scottish health records, enabled individual-level record linkage of nine databases: Community Health Index (CHI) register, NHS GGC Shielding List, Egton Medical Information Systems (EMIS) and Vision, Electronic Communication of Surveillance in Scotland (ECOSS), Prescribing Information System (PIS), Strathclyde Electronic Renal Patient Record (SERPR), Rapid Preliminary Inpatient Data (RAPID), and death certificates.

The CHI register provided sociodemographic information (age, sex, area socioeconomic deprivation). Deprivation was measured using the Scottish Index of Multiple Deprivation (SIMD), derived from seven domains - income, education, health, employment, crime, housing, and access to services – and categorised into general population quintiles. ECOSS collects laboratory data on infectious diseases, including test date and result. Albasoft software extract data from the family practitioner electronic health record systems EMIS and Vision, and PIS collects data on medications prescribed by family practitioners. SERPR records data on renal replacement therapy and transplantation. RAPID collects real-time data on hospitalisation, including dates of admission and discharge, and type of ward, and the Scottish Morbidity Record 01 (SMR01) subsequently records the relevant disease codes. Death certificates provide the date and cause of all deaths, whether in-hospital or in the community. Follow-up data were available until the end of May 2020, before the shielding recommendation was lifted.

Supplementary Table 1 lists the criteria for the shielded and medium risk categories applied at the time of data extraction. All remaining patients are categorised as low-risk. The Scottish list of high-risk individuals is compiled centrally, and regularly updated, using family practitioner, hospital admission, disease registry and medication data. Family practitioners check the completeness and accuracy of the list before letters, recommending shielding, are sent to patients. The NHS GGC Shielding List we used contains the validated data including the criterion satisfied. We ascertained moderate risk individuals using Albasoft extraction of EMIS and Vision data, and PIS data.

Separate models were conducted by overall risk category (low-risk, moderate-risk or shielded) and by the individual criteria for the moderate-risk and shielded categories. The four general population outcomes investigated were: confirmed COVID-19 infection; COVID-19 related hospitalisation; COVID-19 related ICU admission; COVID-19 related mortality. The three outcomes investigated among those with confirmed infection were: COVID-19 related hospitalisation; COVID-19 related ICU admission; and COVID-19 related case fatality.

Laboratory-confirmed cases were defined as positive PCR test. Clinically-confirmed cases were defined as either positive PCR test or death from COVID-19 without testing. COVID-19 related deaths were defined as International Classification of Diseases 10^th^ revision (ICD-10) code U07.1 or U07.2 recorded on the death certificate. COVID-related hospitalisation was defined as an SMR01 hospitalisation record with an ICD code U07.1 or U07.2 or, for more recent admissions, a RAPID hospitalisation record plus positive PCR test taken between two weeks before and two days after hospitalisation. ICU admission during such hospitalisations was assumed to be COVID-related.

Sociodemographic characteristics were compared by risk category using chi-square tests. Poisson regression models with robust standard errors were used to compare risk ratios (RR) for the shielded and moderate-risk categories referent to the low-risk category. The models were run univariately; then adjusted for sex and SIMD quintile as potential confounders. Age was not included as a covariate because it was a moderate-risk criterion. The models were re-run using the individual criteria for the shielded and moderate-risk categories as the exposure variables, referent to the low-risk category.

Population attributable fractions (PAFs) were calculated, from prevalence and adjusted RR, to determine the proportion of each outcome that could be attributed to being shielded and moderate-risk, as well as the proportion due to each individual criterion. The PAFs of individual criteria were proportionally calibrated so that their sum equated to the overall PAF of the relevant risk category. PAF confidence intervals were estimated using bootstrapping (x 1000).

### Ethical approvals

The study was approved by the NHS GGC Primary Care Information Sharing Group and the NHS GGC Local Privacy Advisory Committee (Reference GSH/20RM005) and was covered by the generic Safe Haven Research Ethics Committee approval (GSH20RM005_COVID_Community).

## Data Availability

The dataset supporting the conclusions of this article is available in the Glasgow Safe Haven.

https://www.nhsggc.org.uk/about-us/professional-support-sites/safe-haven/services/

## Acknowledgements

We are extremely grateful to the following for their support and hard work in extracting and linking the datasets and providing access to them: Charlie Mayor (Data Safe Haven Manager), Alison Hamilton (Safe Haven Project Manager), Jonathan Todd (Head of Information Management), Imran Sadat (Data Manager) and Neil Hillen (Data Analyst).

## Contributors

FH and BJ contributed equally and are joint-first authors. BJ and JP conceptualised the study, interpreted the data, and wrote the first draft of the manuscript. FH analysed the data and wrote the first draft of the manuscript. All other authors interpreted the data and critically revised the manuscript. All authors approved the final submitted version of the manuscript. BJ, FH, and JP serve as the guarantor of the manuscript and accepts full responsibility for the work and/or the conduct of the study, had access to the data, and controlled the decision to publish. The corresponding author attests that all listed authors meet authorship criteria and that no others meeting the criteria have been omitted.

