## Supplement for "Comparison of COVID-19 outcomes among shielded and non-shielded populations: A general population cohort study of 1.3 million"

[†]Joint-first author

Bhautesh D Jani PhD
Clinical Senior Lecturer in General Practice and Primary Care
Institute of Health and Wellbeing, University of Glasgow
Glasgow G12 9LX, UK


Frederick K Ho PhD
Research Associate
Institute of Health and Wellbeing, University of Glasgow
Glasgow, G12 8RZ, UK


David J Lowe MSc
Consultant in Emergency Medicine
Queen Elizabeth University Hospital, NHS Greater Glasgow and Clyde
Glasgow, G52 4TF, UK


Jamie P Traynor MD
Consultant Nephrologist
Queen Elizabeth University Hospital, NHS Greater Glasgow and Clyde
Glasgow, G52 4TF, UK


Sean MacBride-Stewart PhD
Lead Pharmacist (Medicines Management Resources)
Pharmacy Services, NHS Greater Glasgow and Clyde
Glasgow, G76 7AT, UK


Patrick B Mark PhD
Professor of Nephrology
Institute of Cardiovascular and Medical Sciences, University of Glasgow
Glasgow, G12 8TA, UK


Frances S Mair MD
Norie Miller Professor of General Practice
Institute of Health and Wellbeing, University of Glasgow
Glasgow G12 9LX, UK

Jill P Pell MD
Henry Mechan Professor of Public Health
Institute of Health and Wellbeing, University of Glasgow
Glasgow, G12 8RZ, UK


***Address for correspondence:**
Professor Jill Pell
Director of the Institute of Health and Wellbeing
University of Glasgow
1 Lilybank Gardens
Glasgow G12 8RZ
United Kingdom


**Supplementary Table 1.** Vulnerable Patient List in UK

| **High Risk** | **Moderate Risk** |
|---|---|
| Severe respiratory disease:<br>• severe COPD<br>• severe asthma (high-dose steroids)<br>• cystic fibrosis | Chronic respiratory disease:<br>• COPD<br>• asthma<br>• emphysema<br>• bronchitis |
| Specific cancers:<br>• Lung cancer plus radical radiotherapy or active chemotherapy<br>• Blood or bone marrow cancers<br>• Bone marrow or stem cell transplant<br> ○ in last 6 months, or<br> ○ still taking immunosuppressants<br>• immunotherapy or continuing antibody treatment<br>• protein kinase inhibitors or PARP inhibitors | Other chronic conditions<br>• heart disease/hypertension<br>• diabetes<br>• kidney disease<br>• liver disease<br>• neurological conditions |
| Pregnant with significant heart disease | Pregnant |
| Immunosuppressive therapy | Weakened immune system due to:<br>• Medical condition<br>• Medication (e.g. oral steroids, chemotherapy) |
| Solid organ transplant | BMI ≥40 kg/$m^2$ |
| Relevant rare diseases and inborn errors of metabolism (e.g. SCID, homozygous sickle cell) | ≥70 years of age |
| Renal dialysis | |

**Supplementary Table 2.** Population attributable fractions for risk categories and risk criteria and COVID-19 outcomes

| | Confirmed COVID-19 | | COVID-19 hospitalisation | | COVID-19 ICU admission | | COVID-19 Death | |
|---|---|---|---|---|---|---|---|---|
| | **%** | **95% CI** | **%** | **95% CI** | **%** | **95% CI** | **%** | **95% CI** |
| ***Moderate*** | | | | | | | | |
| *Overall* | 42.06 | 41.49-42.64 | 53.28 | 52.78-53.78 | 22.96 | 17.70-29.09 | 75.30 | 75.09-75.51 |
| Chronic respiratory disease | 3.56 | 3.29-3.83 | 4.84 | 4.58-5.09 | 0.99 | -2.93-5.28 | 2.23 | 2.00-2.50 |
| Heart disease | 4.83 | 4.73-4.94 | 5.97 | 5.86-6.08 | 1.92 | 0.27-3.55 | 6.19 | 6.10-6.28 |
| Hypertension | 5.33 | 5.19-5.45 | 7.06 | 6.93-7.18 | 10.82 | 9.41-12.11 | 4.99 | 4.90-5.09 |
| Diabetes | 10.39 | 10.17-10.58 | 12.96 | 12.77-13.15 | 9.87 | 7.09-12.63 | 12.00 | 11.82-12.18 |
| Weakened immune system | 0.15 | 0.13-0.16 | 0.26 | 0.24-0.28 | - | - | 0.34 | 0.32-0.36 |
| ≥70 years of age | 17.81 | 17.62-18.01 | 22.19 | 21.98-22.43 | - | - | 49.55 | 49.18-49.94 |
| ***High*** | | | | | | | | |
| *Overall* | 7.62 | 7.45-7.79 | 12.70 | 12.46-12.90 | 2.69 | 1.44-4.10 | 13.22 | 13.02-13.39 |
| Severe respiratory disease | 3.35 | 3.24-3.46 | 6.08 | 5.97-6.20 | - | - | 7.01 | 6.89-7.11 |
| Specific cancers | 0.88 | 0.84-0.92 | 1.42 | 1.37-1.48 | - | - | 1.31 | 1.27-1.36 |
| Immunosuppressive therapy | 1.41 | 1.36-1.47 | 2.43 | 2.36-2.51 | - | - | 2.05 | 1.99-2.11 |
| Solid organ transplant | 0.39 | 0.37-0.42 | 0.73 | 0.68-0.77 | - | - | 0.57 | 0.54-0.60 |
| Rare diseases and IEM | 0.84 | 0.79-0.88 | 1.17 | 1.11-1.23 | - | - | 1.72 | 1.64-1.81 |
| Renal dialysis | 0.75 | 0.68-0.82 | 0.86 | 0.78-0.94 | - | - | 0.55 | 0.50-0.61 |

CI confidence interval; IEM inborn errors of metabolism

**Supplementary Figure 1.** Cumulative population attributable fractions (lines) and population prevalence (bars) for individual risk criteria

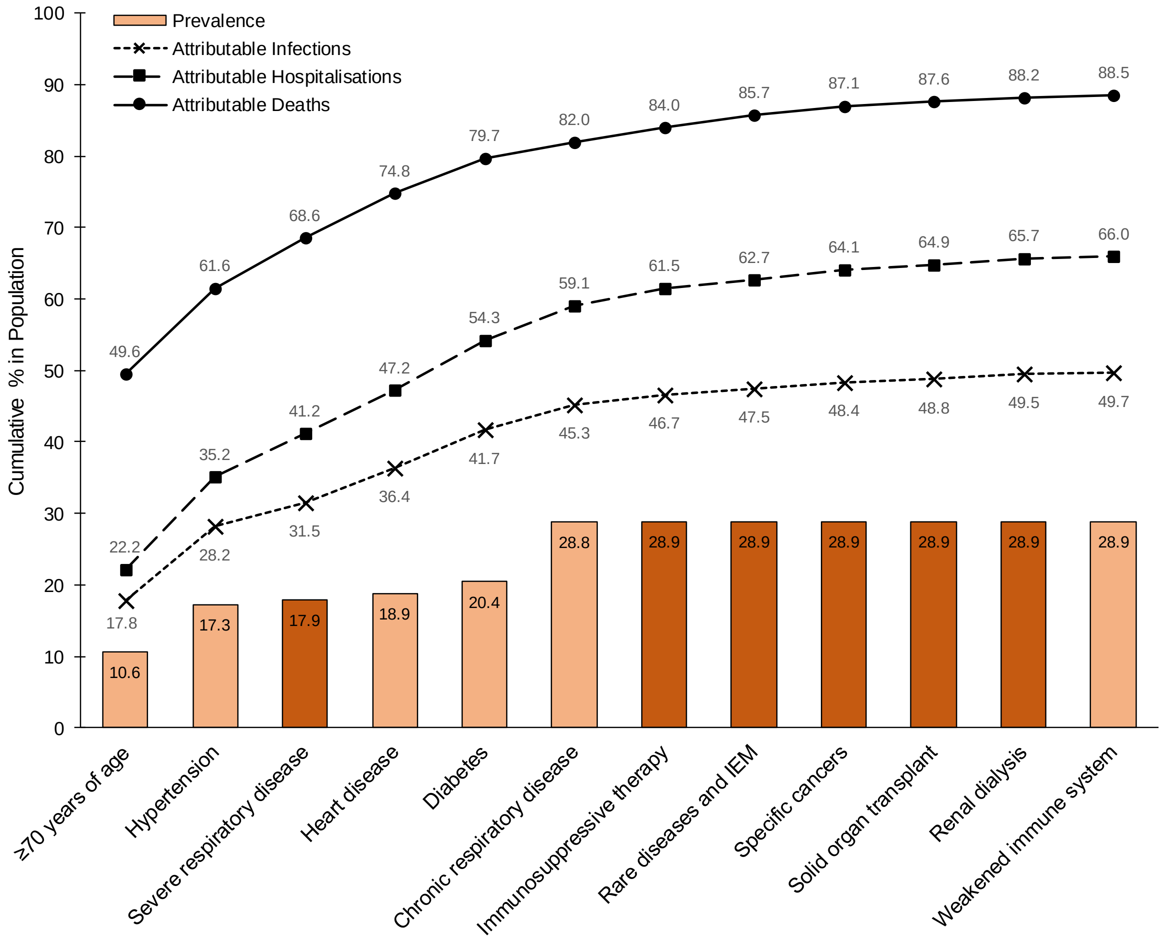


Brown – shielded

Pale orange – moderate-risk
